# CodeMergeR: A shiny-based application for codelist integration

**DOI:** 10.64898/2025.12.07.25341784

**Authors:** Vjola Hoxhaj, Judit Riera-Arnau, Sima Mohammadi, Miriam CJM Sturkenboom, Constanza L Andaur Navarro

**Affiliations:** Department Data Science & Biostatistics, Julius Center for Health Sciences and Primary Care, University Medical Center Utrecht, Utrecht University, Utrecht, The Netherlands; Clinical Pharmacology Service, Vall d’Hebron Hospital Universitari, Vall d’Hebron Barcelona Hospital Campus, Universitat Autònoma de Barcelona, Barcelona, Spain; Vaccine Monitoring Collaboration for Europe (VAC4EU), Brussels, Belgium

**Keywords:** Medical Informatics Applications, Reproducibility, Electronic Health Records, Health Information Interoperability, Software Design, Clinical coding

## Abstract

**Objective:** To describe CodeMergeR, an open-source R shiny application for the standardization, cleaning, and integration of individual concept codelists into a master file for real-world evidence (RWE) studies.

**Material and Methods:** CodeMergeR was designed to align with concept codelists that are outputted by CodeMapper v1.0. The application includes two main modules: (1) Conformance and Coherence, which validates file naming conventions, metadata consistency, and structural integrity; and (2) Cleaning and Standardization, which removes duplicates, corrects formatting issues, and merges validated codelists. Functional and performance testing were conducted using real-world library metadata and codelists obtained from the VAC4EU SharePoint library, including manually introduced errors to assess detection capabilities.

**Results:** The CodeMergeR application successfully identified and resolved a wide range of structural and semantic issues, achieving an overall error detection rate of 96.8% out of 38 errors in the functional testing. The application processed over 50 clinical concept folders and 10 algorithms in under one minute during performance testing. Key issues detected were related to file names, duplications of concepts, formatting anomalies of codes (e.g., scientific notation, rounding), and concept metadata mismatches.

**Discussion:** CodeMergeR shows the need for reproducible and scalable concept codelist management for RWE. In contrast to existing tools such as CodelistGenerator, ATHENA, and OpenCodelists, CodeMergeR fills an important quality aspect in RWE generation and transparency.

**Conclusion:** CodeMergeR contributes to improved reproducibility, scalability, and transparency in individual codelist management in RWE studies.

**KEY POINTS:**

- Health data collected via electronic records (real-world data, RWD) have shown to be a valuable source of information to generate real world evidence (RWE) on effectiveness and safety of medicines and vaccines.
- Very little guidance exists on codelist integration, even though they are essential for semantic harmonization and identification of clinical concepts in RWD. Their manual integration is error-prone and time-consuming, making this an important and under-addressed opportunity to improve RWE quality.
- CodeMergeR is an open-source R Shiny application developed to standardize, verify, and integrate individual concept codelists into a harmonized master file for analytical pipelines in RWE studies.

**PLAIN LANGUAGE SUMMARY:** Researchers who study the real-world use and effects of medicines often use data from electronic health records. This type of data, called real-world data (RWD), is generally analyzed in a federated manner (data stay local) using a common protocol, common analytics and a common data model. In this way RWD from different geographical regions and systems can be used even if diagnoses and treatments are recorded in different vocabularies. To make sure these can be retrieved, compared and combined, researchers use codelists, a set of files with a list of medical codes related to the clinical event of interest. However, concatenation of files can be difficult and time-consuming due to differences in formats, naming styles over time and several people contributing to its creation.

To help with this, we created CodeMergeR, a free, open-source tool that helps researchers clean, check, and combine codelists automatically. It finds and fixes common problems, like missing files, different naming styles, or formatting errors. CodeMergeR fits easily into the tools built on the ConcePTION common data model by the Vaccine Monitoring Collaboration for Europe (VAC4EU) and has been shown to detect almost all issues (96.8%) quickly and accurately.

CodeMergeR helps make studies using codelists more reliable, faster, and easier to repeat. In the future, it could also be used in other research networks that use multiple vocabularies.

## INTRODUCTION

The growing availability of electronic health records (EHR), including medical records, administrative databases (e.g., billing and claims), and disease registries is transforming healthcare research. Collectively known as Real-World Data (RWD), these data sources play an important role in generating real-world evidence (RWE).^1–3^ To transform RWD into RWE, researchers must first define study variables, —that is, specific clinical concepts with roles such as outcome, exposure, or co-variate. These variables are typically retrieved using standardized codes for diagnosis, treatments, procedures, and laboratory results from vocabularies like the International Classification of Diseases (ICD), , Systematized Nomenclature of Medicine Clinical Terms (SNOMED CT), Read Codes, and International Classification of Primary Care (ICPC), among others.^4–7^

A codelist is a curated set of codes that represents a clinical concept across different vocabularies. For example, Guillan-Barré Syndrome may be recorded in ICPC with N94.01, in SNOMED CT with 40956001, and in ICD-10-CM with G61.0.^6,8,9^ Codelists are essential for semantic harmonization in RWE, as they enable automated and transparent code retrieval across data sources.^10^ However, creating and updating codelists remains an important challenge due to dynamic nature of vocabularies which require frequent updates.

Research networks have taken different approaches towards semantic harmonization. The Observational Health Data Sciences and Informatics (OHDSI) network, for example, requires translation of all original codes into a common vocabulary and uses ATLAS to compile concepts. The CodelistGenerator from DARWIN EU supports the systematic extraction and aggregation of codelists for vocabularies in the Observational Medical Outcomes Partnership (OMOP) Common Data Model (CDM). VAC4EU and EU PE&PV research networks use the ConcePTION CDM, which eliminates the need for upfront vocabulary translation by the data holder.^11^ Instead, it performs vocabulary mapping within the study scripts, for speed, transparency and flexibility.

Currently, researchers often create individual codelists stored in various formats such as spreadsheets or text files, without consistent naming conventions or standardized format structures. As highlighted by Matthewman et al., the use of metadata and clear, consistent naming conventions is essential for ensuring codelist’s transparency, minimizing errors, and promoting reuse in future studies.^10^ Within VAC4EU, a harmonized approach is implemented: codelists are first created by a coordinator using CodeMapper and then tagged by medical reviewers using specific labels to indicate the level of specificity for each code in relation to the study objective.^12^ However, creating, tagging, and maintaining codelists with multiple reviewers requires substantial effort and oversight to ensure high-quality codelists. The process of merging individual concept codelists into a single master study codelist is error-prone, as it can lead to issues such as code duplication, formatting inconsistencies, and accidental omission of relevant concepts.

In this article, we introduce CodeMergeR, an open-source R shiny application that provides an efficient and user-friendly tool to support researchers in the standardization, cleaning, and integration of individual concept codelists into a harmonized study codelist for RWE studies.

## 2. METHODS

### 2.1 File workflow

CodeMergeR is currently designed to align with a prespecified folder and file structure used in VAC4EU. VAC4EU has already adopted many of the indicators explained by *Matthewman et al.*, such as metadata, clinical concept definition, review and publication.^10^ A dedicated taskforce composed by a coordinator (SM) and medical doctors is responsible for creating and reviewing collaborative codelists files obtained from CodeMapper v1.0, which maps clinical concepts across multiple vocabularies based on the Unified Medical Language System (UMLS).^12^ If available, local vocabularies (i.e., not necessarily standardized internationally) are provided by collaborators and manually mapped to the corresponding clinical concepts.

Usually, researchers need hundreds of individual codelists for specific clinical concepts relevant to their study. After creation of codelists in CodeMapper v1.0, these files were uploaded to Sharepoint within a nested folder structure. All folders followed standardized naming conventions. The folder name comprises four elements: first, a letter indicating the system related to the codelists (e.g., “E” for Endocrine, metabolic, or nutritional diseases or “I” for Infectious diseases); second, an abbreviation representing the concept; third, the type of variable for which the concept is made, such as “COV” for covariate or “AESI” for outcomes, and fourth, the full name of the codelist. These four elements are separated by underscores, resulting in three underscores in total within the folder name. The name of the file (codelist) containing the diagnostic codes is derived from the folder name, except for the full name of the concept. This combination is referred to as the variable_name. A worksheet with the same variable_name must be present. The file format extension *.xls or *.xlsx was chosen for uploading due to its user-friendly interface, enabling the medical team to store comments and highlight specific cells for discussion and approval.

#### 2.1.3 Mandatory columns in working files

Each codelist file was required to have a set of mandatory columns which will be used to create a master file (Box 1). These columns are different for the files in the Algorithms/ folder and the ones in the clinical concept folders (Box 2). Examples can be found in the Supplemental File 1.

**Box 1:** Mandatory fields or columns for the file in each clinical concept folders.

| Fields* | Definition | Example |
| --- | --- | --- |
| **Coding system** | Standardized vocabulary used to classify medical concepts, such as diseases, procedures, medications, and clinical events, through unique codes | ICD10 |
| **Code** | Unique identifier, typically alphanumeric, assigned to represent a specific medical concept | E10 |
| **Code name** | Label or description assigned to a specific set of medical codes representing a condition, procedure, or medication. | brittle diabetes (mellitus) |
| **Concept** | Unique, unified representation of a medical or health-related concept that links equivalent terms, codes, and descriptions from different vocabularies and coding systems based on UMLS (i.e., concept unique identifier CUI). | C0011854 |
| **Concept name** | UMLS based descriptive term that represents a unique clinical or health-related concept and serves as a label for the concept. | Diabetes Mellitus, Insulin-Dependent |
| **Tags** | Level of specificity used when selecting codes to represent a medical concept | narrow |

*Letter case and spelling is important when running the app.*
*\*These fields are retrieved from the CodeMapper v1.0 tool.*

**Box 2:**
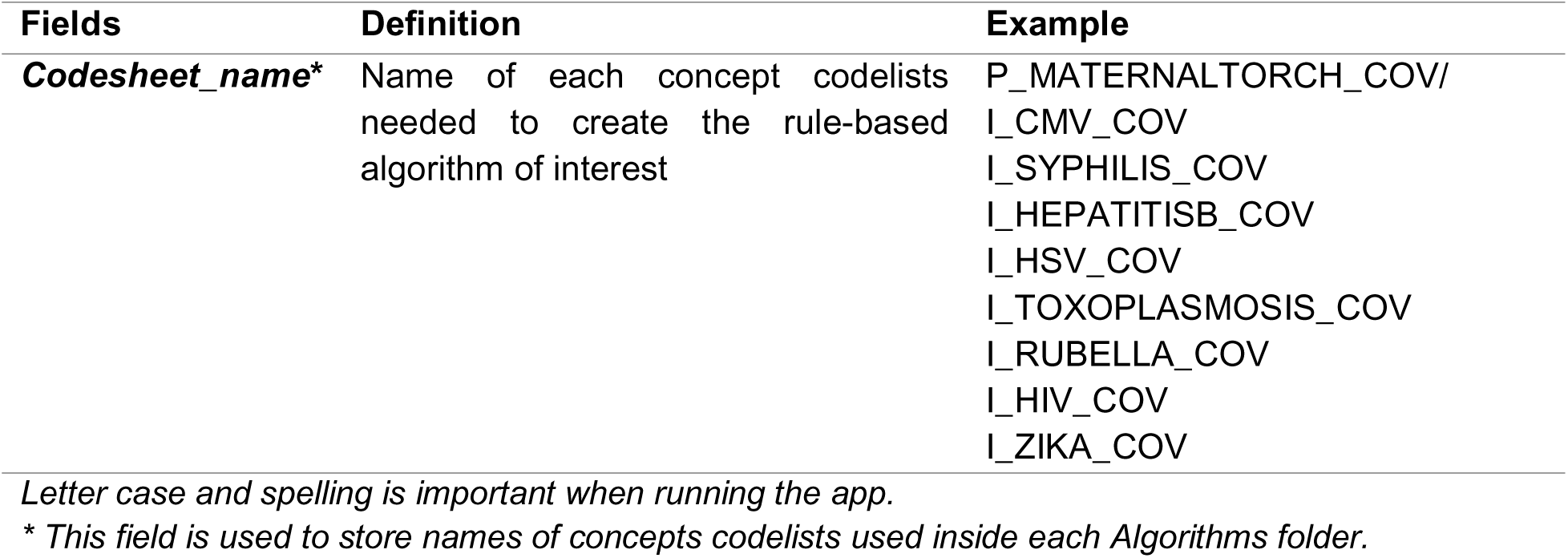
Mandatory fields or columns for the file in Algorithms/ folders Fields Definition Example.

Box 2: Mandatory fields or columns for the file in Algorithms/ folders
| Fields | Definition | Example |
| --- | --- | --- |
| ***Codesheet_name\**** | Name of each concept codelists needed to create the rule-based algorithm of interest | P_MATERNALTORCH_COV/<br>I_CMV_COV<br>I_SYPHILIS_COV<br>I_HEPATITISB_COV<br>I_HSV_COV<br>I_TOXOPLASMOSIS_COV<br>I_RUBELLA_COV<br>I_HIV_COV<br>I_ZIKA_COV |

*Letter case and spelling is important when running the app.*
*\* This field is used to store names of concepts codelists used inside each Algorithms folder.*

#### 2.1.4 Library metadata

The library metadata file consists of several worksheets that document information about codelists for various clinical concepts, such as concepts, procedures, and medicines. It serves as a central repository to store information about existing codelists, review status, and use of codelist across studies, as well as codelists to be created. The worksheet particularly relevant for CodeMergeR is called *CDM_Events* which focuses on algorithms and clinical concepts, and contains key fields that facilitate the identification, creation and processing (i.e., validation and combination) of codelists (Box 3).

**Box 3:** Mandatory fields or columns in the excel worksheet CDM_Events.

**Box 3: Mandatory fields or columns in the excel worksheet CDM\_Events**
| Fields* | Definition | Example |
| --- | --- | --- |
| **Event_abbreviation/**<br>**Variable Name** | Represents the *variable_name* derived from the folder and file naming convention. | C_AMI_AESI_Acute myocardial Infarction |
| **Covariates with No Code** | Identifies concepts for which codelists are not yet available (i.e., an Excel file is not present in the corresponding folder), those planned for future development, or those that have been archived and are no longer in use. | Yes – When codelist is to be created.<br>(empty) – When codelist is available. |
| **Study Name** | Indicates the studies requiring specific Algorithms or CONCEPTSs, enabling filtering of the codelist library metadata based on the needs of a particular research project. | COVID19_AESI |
| **Type of Events** | Specifies the classification of the event | AESI |

*AESI* – Adverse Event of Special Interest; *COV* – Covariate, *PrA* – Codes used in the pregnancy algorithm developed within ConcePTION.<sup>13</sup> (\*) While these administrative columns are valuable for broader codelist management, they are not directly utilized by CodeMergeR.

### 2.2 CodeMergeR

#### 2.2.1 Design & Architecture

We aimed to develop a tool that incorporates automated checks to detect invalid folder names and flag inconsistencies within independent codelist files. The design of CodeMergeR consists of four components: input files, a user-friendly front-end interface, a back-end processing dataflow, and export files.

In Figure 1, the app workflow begins with the input and import phase, where users can upload multiple Excel files containing independent codelists and the library metadata, and specify the study name. The conformance and coherence module which is part of the back-end processing ensures file compliance with predefined requirements, such as the number and type of files, file extensions, and naming conventions, while also verifying consistency across library metadata, algorithm folders, and clinical concepts. Next, the Cleaning and Standardization module identifies and removes duplicate entries, corrects formatting issues, and eliminates invalid characters from key fields such as code and vocabulary. This results in a curated, standardized codelist ready to be exported. The specific checks performed by CodeMergeR in both modules are described in Boxes 4 and 5. Furthermore, the export functionality enables users to download the master file (i.e. the concatenated study codelist) and the pregnancy algorithm (PrA) codelist in CSV format. Also, files reporting errors (when applicable) and two summary tables of included concepts, vocabularies and tags (tags_details.csv and summary.csv). All export files are saved locally within the ‘cleaned_codelist” folder within the app main directory, which is the local folder where the code of the app is saved.

**Figure 1.**
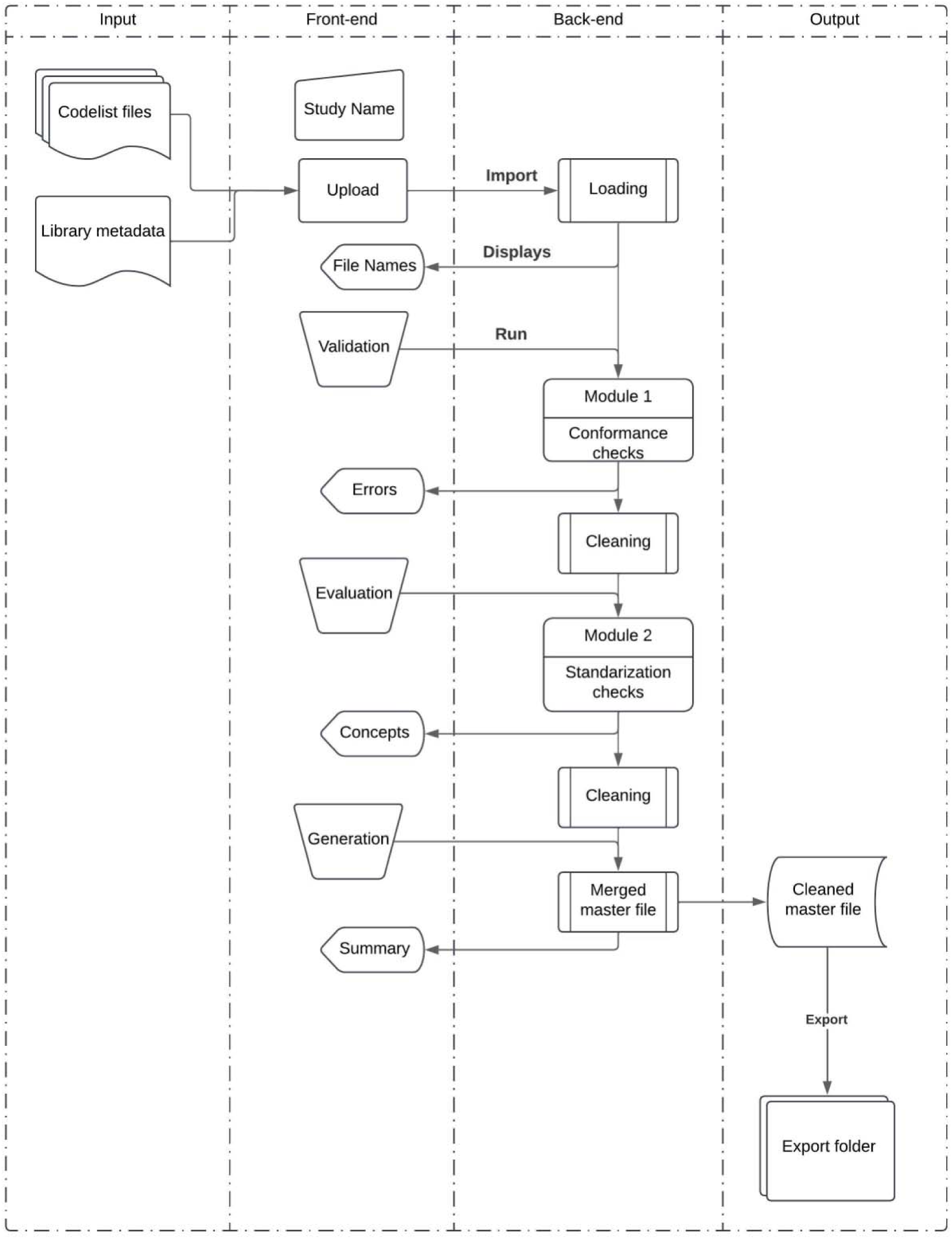
CodeMergeR. The process begins with the input of codelist files and the library metadata file. The front-end interface allows users to specify the study name, upload files, and view validation results. The back-end processing includes two modules: Module 1 performs conformance checks to validate folder and file structure, while Module 2 conducts standardization checks to clean and harmonize the data. The final output includes a cleaned master codelist and associated summary files, which are exported for use in RWE studies.

Internal validity checks are included through various steps of the app to ensure input files conform to expected standards. For example, if the library metadata file contains all required fields. Details of each check are provided in Box 6.

#### 2.2.2 User-interface and usability features

CodeMergeR starts with a pop-up message when initializing the app, which describes a summary of the steps within the app. After, two tabs open, one for module Conformance and Coherence (tab 1) and one for module Cleaning and Standardization (tab 2). Tab 1 displays information about the uploaded data folder, including information such as the file name, date of execution, and the results of each individual check in module conformance and coherence. A box explaining all rules and guidelines on folder structure and naming (conformance) and folder availability (coherence) is also present in the first tab. A clickable legend box is available for each tab where users can personalize which checks they would like to implement.

Tab 2 displays the summary of all included concepts and tags together with their corresponding vocabularies, and the results of each individual check in module cleaning and standardization. Each tab has an option to capture and save screenshots for documentation purposes. We have made available a short video in our repository for visualization (https://github.com/vjolahoxhaj/CodeMergeR).

**Box 4:** Module 1 Conformance and Coherence.

**Box 4: Module 1 Conformance and Coherence**
| Check type | Description | System Behavior |
| --- | --- | --- |
| **#1 Folder Names** | Checks whether all folder names have 3 underscores. | Valid folders are copied into a newly created clinical concepts folder named Conceptsets/Algorithms within the app directory. Erroneous folders are excluded from further processing. |
| **Displayed messages\*\*** |  |  |
| List of erroneously named folders |  |  |
| "All folders' names in clinical concepts are correctly formatted" |  |  |
| "Concept folder name does not have at least three elements" |  |  |
| **#2 Files with no codes** | Checks for presence of Excel file (.xls or .xlsx) inside the concept folder for those folders that have been specified as 'Covariates with no codes' in the library metadata file. | Erroneous folders are removed from further processing.<br>List of erroneous folders. |
| **Displayed messages\*\*** |  |  |
| "This folder is marked as 'Concept with no codes' but contains an Excel file, which is not expected. This folder will be removed" |  |  |
| "All 'Concepts with no codes' folders are correctly specified and contain no Excel files" |  |  |
| **#3 Study name presence\*** | Checks if the name of the study in the app is present as a field name in the library metadata file | App stops |
| **Displayed messages\*\*** |  |  |
| "The study name you entered is found as a column in library metadata" |  |  |
| "The study name you entered is not found in the library metadata. Please check for typos or column naming" |  |  |
| **#4 Excel File Count** | Check if only one Excel file (.xls or .xlsx) is present inside the concepts folder | Erroneous folders are removed from further processing.<br>List of erroneous folders. |
| **Displayed messages\*\*** |  |  |
| "All concepts' folders contain exactly one Excel file" |  |  |
| "The following folders contain zero Excel files. Create the clinical concept Excel file" |  |  |
| "The following folders contain multiple Excel files. Keep only one Excel file in case of multiple files" |  |  |
| **#5 Folder and Excel File Name Match** | Check if the name of the concept folder and the Excel file inside match | Erroneous folders are removed from further processing.<br>List of erroneous folders. |
| **Displayed messages\*\*** |  |  |
| "The following folders have mismatched names between the folder and the Excel file. Match the name of the Concept folder and Excel file located inside the folder" |  |  |
| "All folders and their Excel files are correctly named and match" |  |  |
| **#6 Excel worksheet Name Match** | Check if the worksheet inside the Excel file has the same name as the file itself | Erroneous folders are removed from further processing.<br>List of erroneous folders. |
| **Displayed messages\*\*** |  |  |
| "The following folders have mismatched names between the Excel file and the worksheet inside. Match the name of Excel file and at least one worksheet of inside this Excel file"<br>"All Excel files contain a worksheet name that matches the file name" |  |  |
| **#7 Mandatory Columns** | Check if all mandatory columns are present within Excel files | Erroneous folders are removed from further processing.<br>List of erroneous folders. |
| **Displayed messages\*\*** |  |  |
| "All Excel files in Concepts folders contain the required columns"<br>"Missing mandatory columns: [columns' names]. Review requirements and add the necessary column names" |  |  |
| **#8 Concepts and Algorithms Match** | Check if all concepts necessary to create the Algorithms exist in the Concepts folder. | Erroneous folders are removed from further processing.<br>List of erroneous folders. |
| **Displayed messages\*\*** |  |  |
| "The following folders are present in *Algorithms* but missing from *Concepts*: [folder names]. Create the necessary Concepts folder for each Algorithm or tag them to the 'Covariates with no codes' column"<br>"All folder names are consistently present in both *Algorithms* and *Concepts*" |  |  |
| *Not applicable for either concepts or algorithms/ folder |  |  |
| **Same displayed messages but adapted to Algorithms/ folder context. |  |  |

**Box 5:**
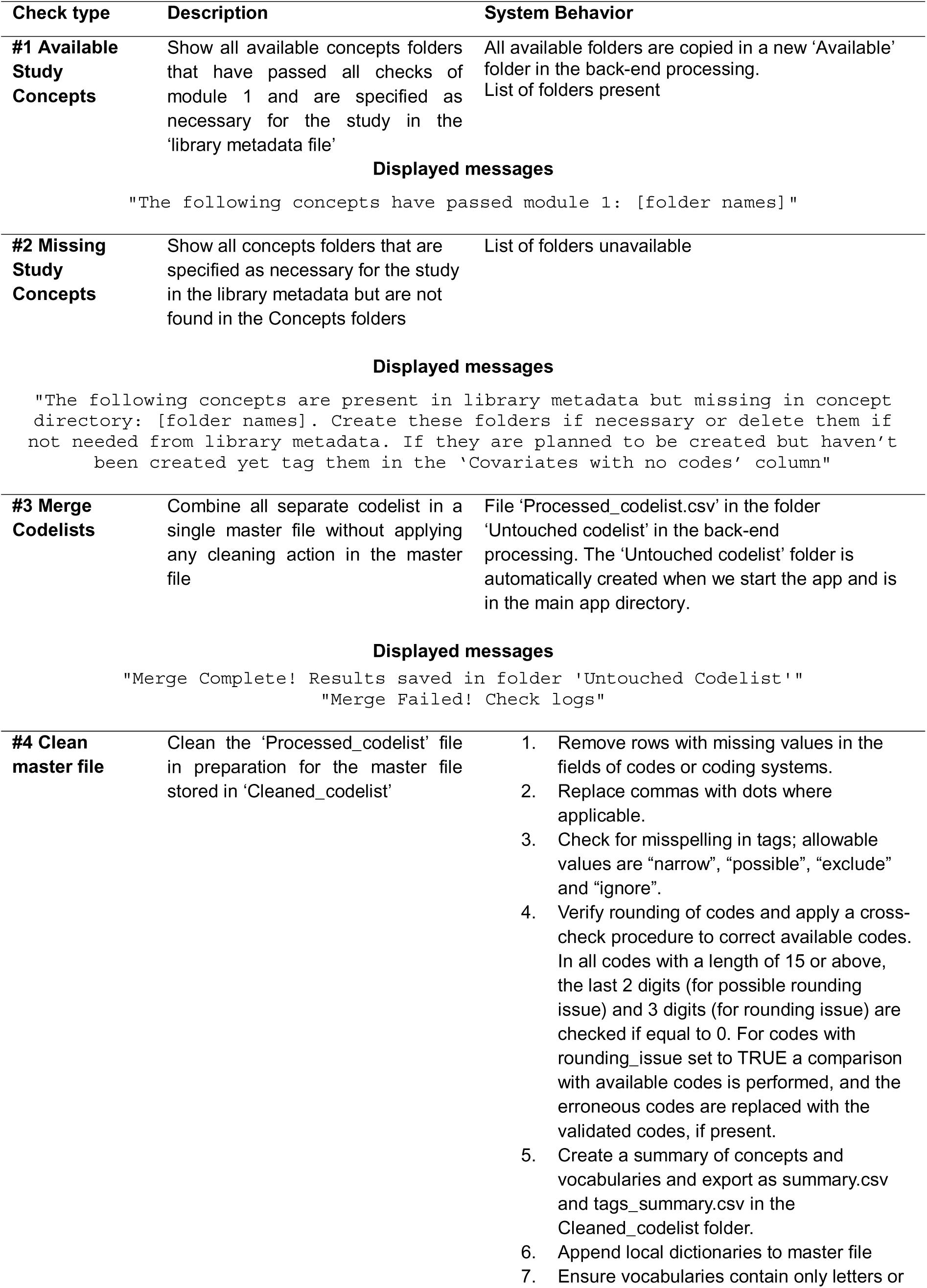
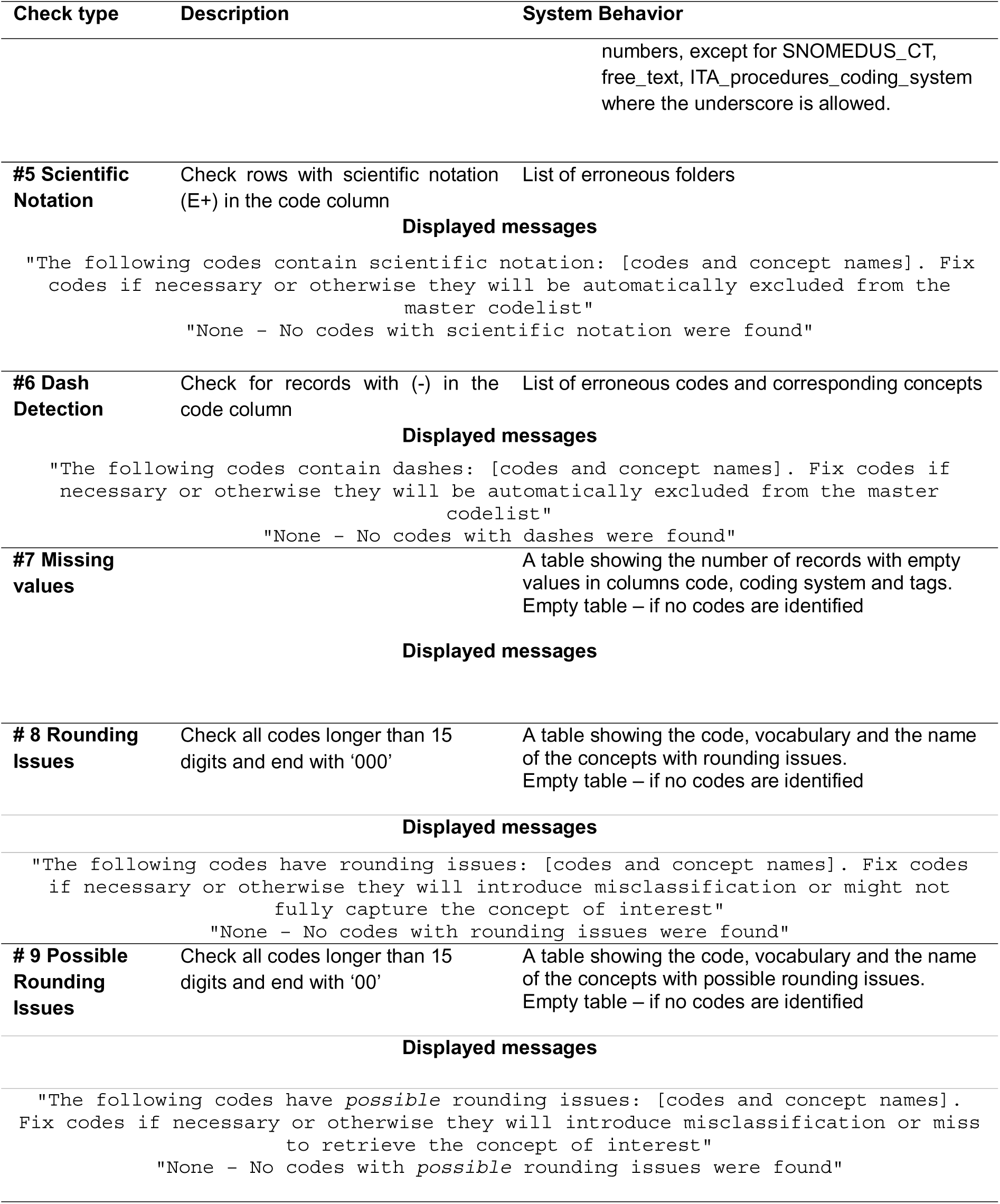
Cleaning and standardization.

**Box 5: Cleaning and standardization**
| Check type | Description | System Behavior |
| --- | --- | --- |
| **#1 Available Study Concepts** | Show all available concepts folders that have passed all checks of module 1 and are specified as necessary for the study in the 'library metadata file' | All available folders are copied in a new 'Available' folder in the back-end processing.<br>List of folders present |
| **Displayed messages** |  |  |
| `"The following concepts have passed module 1: [folder names]"` |  |  |
| **#2 Missing Study Concepts** | Show all concepts folders that are specified as necessary for the study in the library metadata but are not found in the Concepts folders | List of folders unavailable |
| **Displayed messages** |  |  |
| `"The following concepts are present in library metadata but missing in concept directory: [folder names]. Create these folders if necessary or delete them if not needed from library metadata. If they are planned to be created but haven't been created yet tag them in the 'Covariates with no codes' column"` |  |  |
| **#3 Merge Codelists** | Combine all separate codelist in a single master file without applying any cleaning action in the master file | File 'Processed_codelist.csv' in the folder 'Untouched codelist' in the back-end processing. The 'Untouched codelist' folder is automatically created when we start the app and is in the main app directory. |
| **Displayed messages** |  |  |
| `"Merge Complete! Results saved in folder 'Untouched Codelist'"`<br>`"Merge Failed! Check logs"` |  |  |
| **#4 Clean master file** | Clean the 'Processed_codelist' file in preparation for the master file stored in 'Cleaned_codelist' | 1. Remove rows with missing values in the fields of codes or coding systems.<br>2. Replace commas with dots where applicable.<br>3. Check for misspelling in tags; allowable values are "narrow", "possible", "exclude" and "ignore".<br>4. Verify rounding of codes and apply a cross-check procedure to correct available codes. In all codes with a length of 15 or above, the last 2 digits (for possible rounding issue) and 3 digits (for rounding issue) are checked if equal to 0. For codes with rounding_issue set to TRUE a comparison with available codes is performed, and the erroneous codes are replaced with the validated codes, if present.<br>5. Create a summary of concepts and vocabularies and export as summary.csv and tags_summary.csv in the Cleaned_codelist folder.<br>6. Append local dictionaries to master file<br>7. Ensure vocabularies contain only letters or |
|  |  | numbers, except for SNOMEDUS_CT, free_text, ITA_procedures_coding_system where the underscore is allowed. |
| **#5 Scientific Notation** | Check rows with scientific notation (E+) in the code column | List of erroneous folders |
| **Displayed messages** |  |  |
| "The following codes contain scientific notation: [codes and concept names]. Fix codes if necessary or otherwise they will be automatically excluded from the master codelist"<br>"None - No codes with scientific notation were found" |  |  |
| **#6 Dash Detection** | Check for records with (-) in the code column | List of erroneous codes and corresponding concepts |
| **Displayed messages** |  |  |
| "The following codes contain dashes: [codes and concept names]. Fix codes if necessary or otherwise they will be automatically excluded from the master codelist"<br>"None - No codes with dashes were found" |  |  |
| **#7 Missing values** |  | A table showing the number of records with empty values in columns code, coding system and tags.<br>Empty table – if no codes are identified |
| **Displayed messages** |  |  |
| **# 8 Rounding Issues** | Check all codes longer than 15 digits and end with '000' | A table showing the code, vocabulary and the name of the concepts with rounding issues.<br>Empty table – if no codes are identified |
| **Displayed messages** |  |  |
| "The following codes have rounding issues: [codes and concept names]. Fix codes if necessary or otherwise they will introduce misclassification or might not fully capture the concept of interest"<br>"None - No codes with rounding issues were found" |  |  |
| **# 9 Possible Rounding Issues** | Check all codes longer than 15 digits and end with '00' | A table showing the code, vocabulary and the name of the concepts with possible rounding issues.<br>Empty table – if no codes are identified |
| **Displayed messages** |  |  |
| "The following codes have *possible* rounding issues: [codes and concept names]. Fix codes if necessary or otherwise they will introduce misclassification or miss to retrieve the concept of interest"<br>"None - No codes with *possible* rounding issues were found" |  |  |

**Box 6:** Internal Validity Checks Implemented in CodeMergeR.

Box 6. Internal Validity Checks Implemented in CodeMergeR
| Validation type | Description | System Behavior |
| --- | --- | --- |
| **Library metadata presence** | Check that the library metadata file can be loaded correctly | App stops |
| **Displayed messages** |  |  |
| "Error reading the library metadata. Please check the file format or permissions" |  |  |
| **Concepts folder presence** | Check for presence of at least one Concepts folder in the uploaded data | App skips all checks |
| **Displayed messages** |  |  |
| "No folders are found in the directory to check" |  |  |
| **Concepts folder – Library metadata comparison** | Verifies if all required folders according to the library metadata file are present in the concept folder directory | Displays error |
| **Displayed messages** |  |  |
| "These Concepts folders are missing but present in the library metadata. Please create it and re-upload before running this check" |  |  |
| **Codelist metadata** | Verifies the structure and content of the library metadata file. | Missing worksheet *CDM_Events*; Displays error |
| **Displayed messages** |  |  |
| "Required worksheet not found in the library metadata. Review requirements and add the necessary worksheet" |  |  |
| **Library metadata file structure** | Checks for missing required fields in the library metadata file. | Displays error |
| **Displayed messages** |  |  |
| "Required columns ['Event_abbreviation / Variable Name', 'Covariates with no codes'] not found in the library metadata. Review requirements and add the necessary columns" |  |  |
| **File format** | Ensures that only valid file types are uploaded. For example, only .zip files. | Displays error |
| **Displayed messages** |  |  |
| "Only .zip files accepted in *Upload Codelist Folder* box" |  |  |
| **Folder availability** | Checks for required folders before performing naming convention checks. | Displays error; List missing folders |
| **Displayed messages** |  |  |
| "No folders are found in directory to check" |  |  |
| "The Concepts folder is missing. Please ensure it exists before running the check" |  |  |

#### 2.2.3 Software

We developed CodeMergeR using R version 4.2.3 within the RStudio environment.^14^ To improve usability, embedded JavaScript was used for dynamic UI components. A mandatory pop-up ensures users read and acknowledge key information by clicking ‘OK’ before use. The app integrates several R packages including shiny, DT, readxl, and tidyverse, and was tested on Windows and macOS.^15–26^ Deployment was carried out via shinyapps.io, ensuring accessibility for end-users without requiring local installation. The source code is maintained in a public repository (https://github.com/vjolahoxhaj/CodeMergeR) accompanied by user documentation to support reproducibility and adoption.

#### 2.2.4 App testing

We conducted functional and performance testing to ensure that CodeMergeR performed as intended using real-world library metadata and codelists available in the VAC4EU library. We also tested edge cases, such as missing or incorrect input files, to confirm that the app handles errors appropriately.

For functional testing, we randomly selected ten clinical concepts in the library metadata and set them to “Yes”: B_ANAEMIA_COV (Anaemia), B_COAGDEF_COV (Coagulation, deficiencies), C_AMI_AESI (Myocardial infarction), C_HF_AESI (Heart failure), E_GOUT_COV (Gout), Im_ANAPHYLAX_COV (Anaphylaxis history), M_ARTRHEU_COV (Rheumatoid arthritis), N_CONVULSION_AESI (Generalized convulsion), N_EPILEPSY_COV (Epilepsy), and Onc_BREASTCANCER_COV (Breast cancer). This allowed us to verify that the app correctly processed selected clinical concepts.

For performance testing, we introduced a mix of correctly structured and manually flawed files in a copy of the VAC4EU library, to simulate common errors encountered in practice, such as incorrect folder naming, invalid code formats, missing files, and inconsistent metadata. An overview of the errors introduced is documented in Supplemental File 2. We evaluated the correctness of the cleaning and integration processes by comparing the automated error outputs with manual review.

## 3. RESULTS

### 3.1 Use case

#### 3.1.1 Installation Requirements and Environment

The app package was obtained from the corresponding GitHub repository and stored locally within the working environment. Detailed instructions for installation and execution are available in the repository. The preparation of the input files involved two components: (i) a .zip file containing the *Concepts* directory, and (ii) library metadata. The *Concepts* directory comprised an *Algorithms* subfolder, containing separate algorithm folders, along with distinct folders for the individual codelists. Within the library metadata, a new column was created to specify the study name. Concepts and algorithms relevant to the study were marked as “yes” in this column. After input configuration, the torun.R script was executed and, a web interface automatically opened in the default browser, guiding the user through the subsequent steps via an on-screen pop-up window.

#### 3.1.2 App testing: features

For functional testing, we deployed CodeMergeR using the tailored library metadata with 10 concepts as required for the study and the full list of independent codelists available in the VAC4EU library. The app correctly processed selected clinical concepts reporting issues in two checks from module Conformance and Coherence and four in module Standardization and Cleaning.

In Module 1, the first issue, check #4: Excel File Count, occurred in B_COAGDIS_AESI_Coagulation disorders all, where multiple Excel files were detected in the folder instead of the expected single file (Figure 2). The second issue, identified in Check #8: Not all necessary Concepts for Algorithm creation are available: C_ARRH_AESI and R_ASTHMAEXACERBATION_AES, was caused by the requirement of these two codelists for the creation of Algorithms present in the main directory (Figure 3).

**Figure 2.**
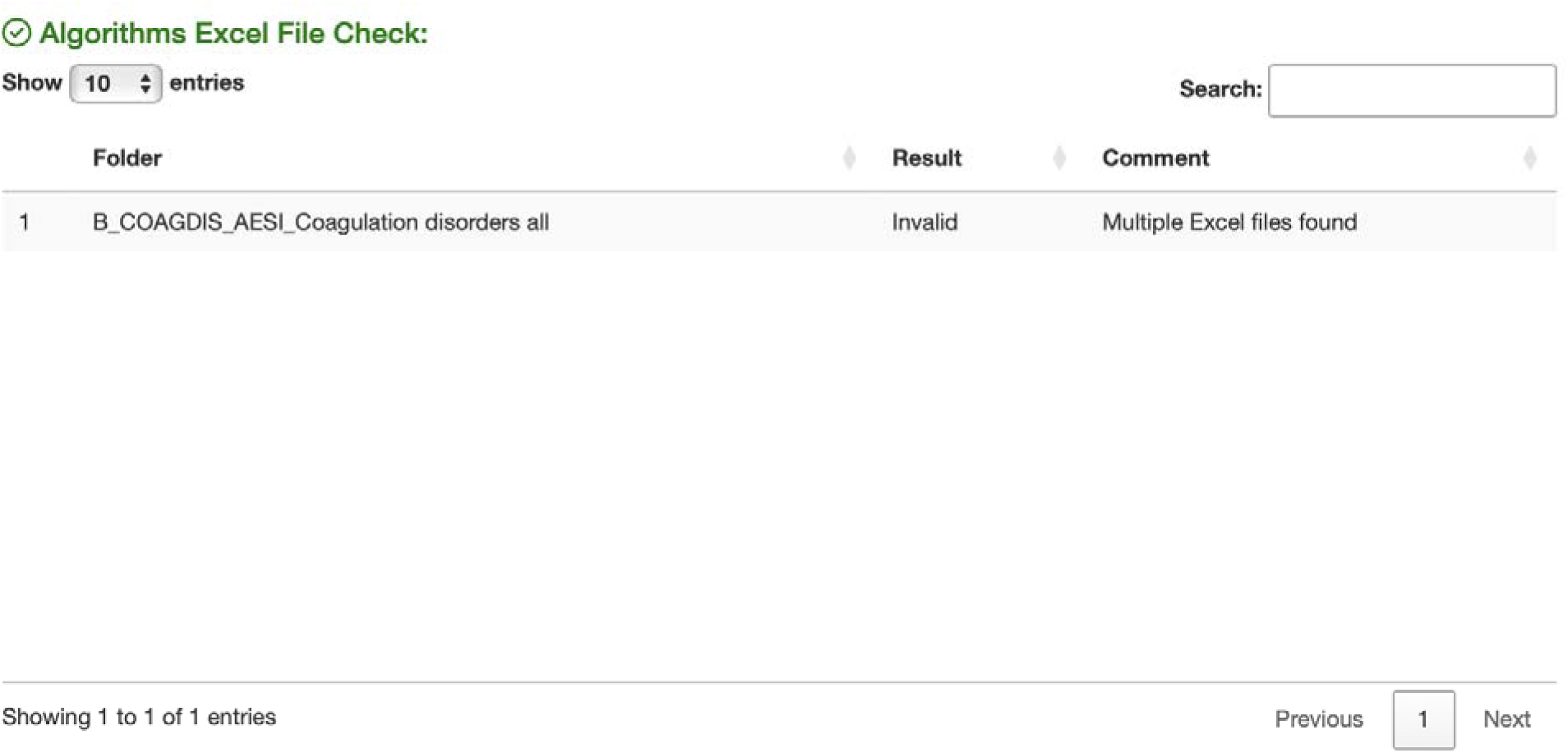
Visualization module 1 check #4. The folder failed the check due to multiple Excel files inside, resulting in an “Invalid” status.

**Figure 3.**
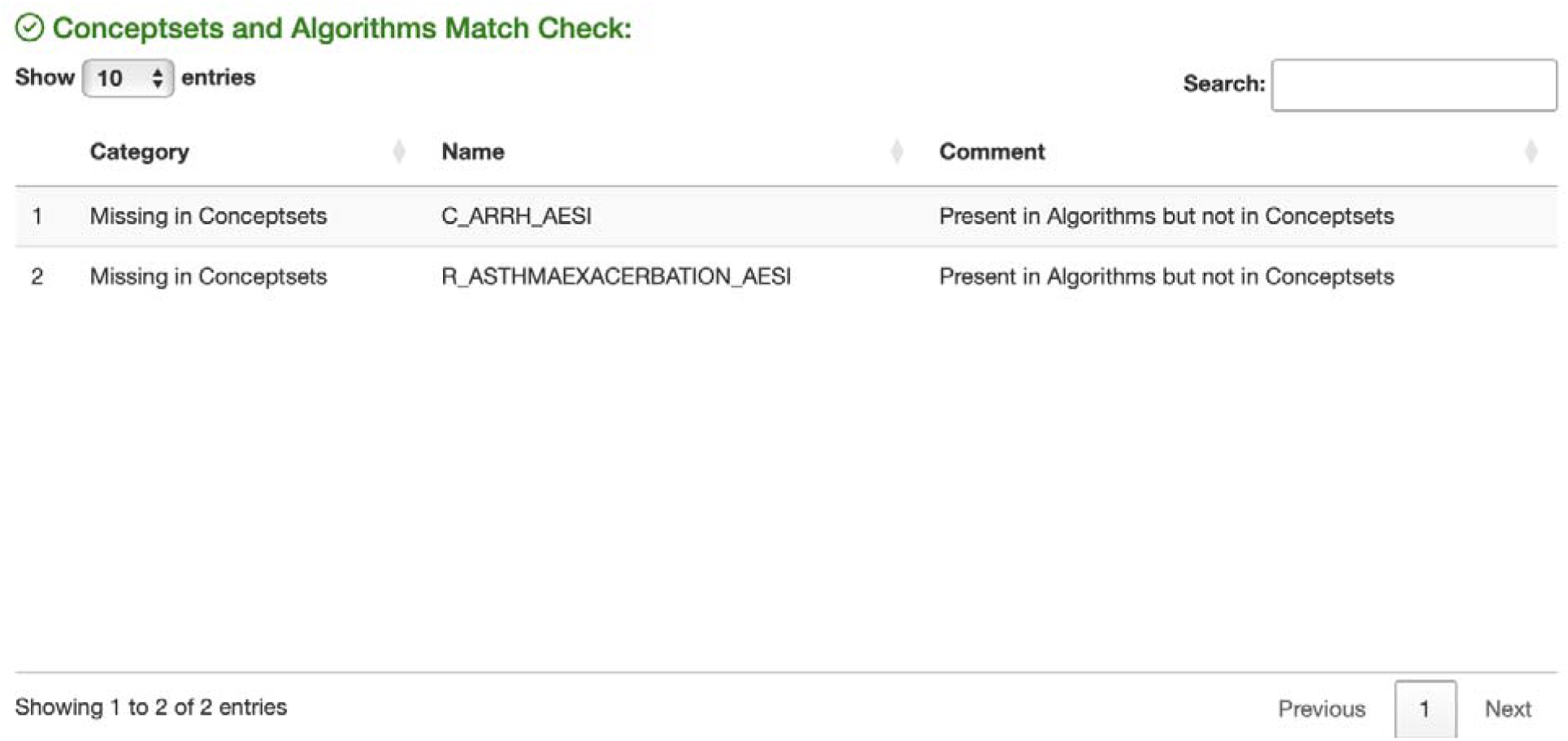
Visualization module 1 check #6. The folder failed the check due to a mismatch or formatting issue in the Excel file, resulting in an “Invalid” status.

In Module 2, the first issue, check #1: Missing Study Concepts, involved B_ANAEMIA_COV being listed as missing (Figure S1). Additionally, eleven structural issues were detected in Check #6: Dash Detection (Figure 4) and four issues in Check #7: Missing Values, all related to pregnancy clinical concepts labeled as PrA (Figure S2). Under Check #9: Possible Rounding, 60 codes were flagged with potential rounding issues, including E_GOUT_COV (n=8), Im_ANAPHYLAX_COV (n=12), N_CONVULSION_AESI (n=1), Onc_BREASTCANCER_COV (n=10), and pregnancy-related clinical concepts (n=29) (Figure S3).

**Figure 4.**
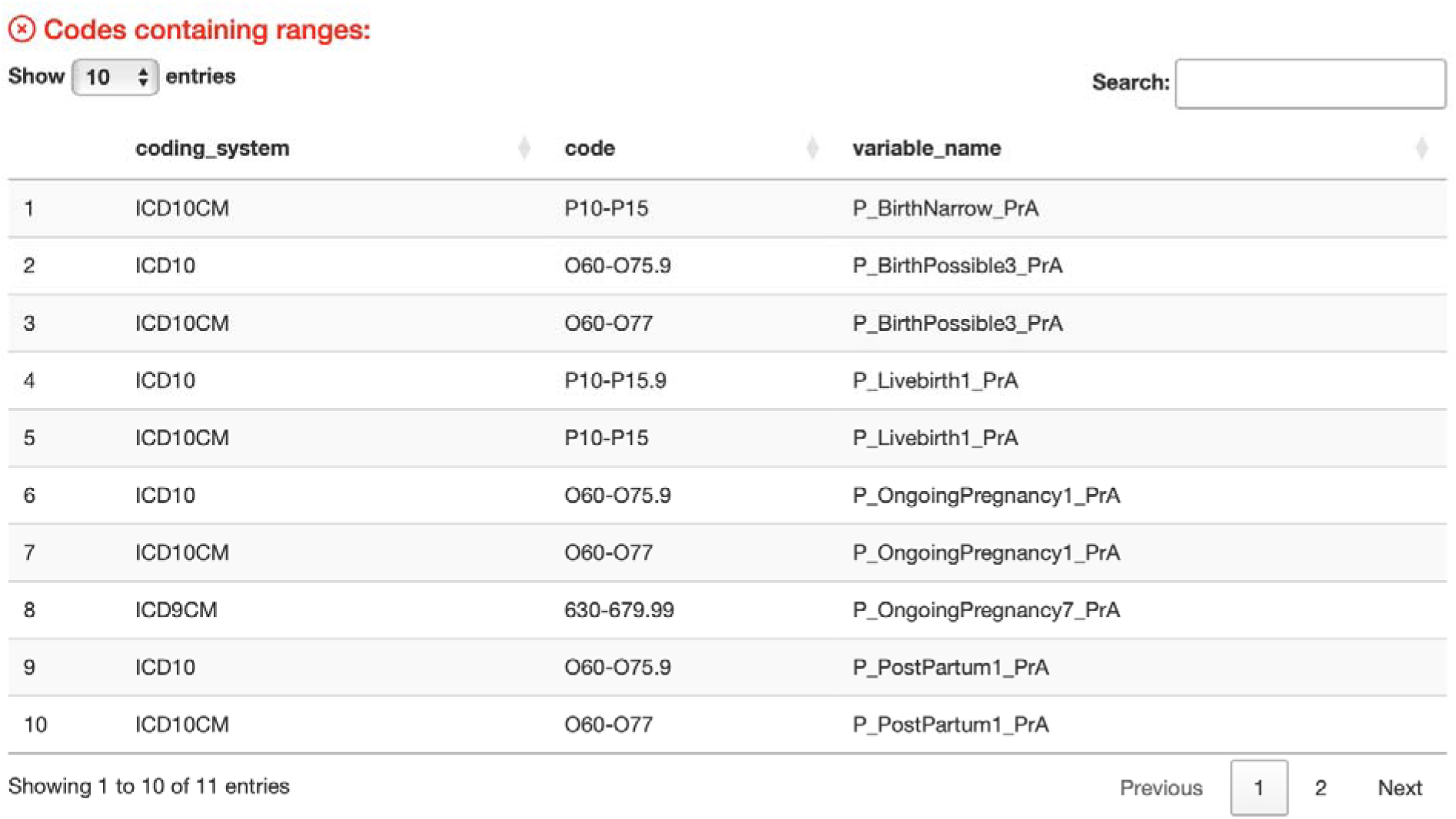
Visualization module 2 check #6. List of diagnostic codes containing range-style values (dash) across different coding systems (ICD-10, ICD-10-CM, ICD-9-CM) used in pregnancy-related clinical concepts. These range-style codes require special handling during validation and standardization to ensure accurate code representation.

#### 3.1.2 App testing: performance

For performance testing, we deployed CodeMergeR using the modified versions of library metadata and 50 concept codelists and 10 algorithms from the VAC4EU library in which errors were introduced. This process ended within one minute. Table 1 we summarize the types and number of introduced errors correctly identified by CodeMergeR. In total, 63 distinct errors were introduced for testing, of which 61 were successfully detected, yielding an overall detection rate of 96.8%. Screenshot of the errors is available in Supplemental File 3.

**Table 1.** Summary of introduced vs. detected errors.

| **Error** | **Examples in test set** | **Checks (Figure)** | **Detected errors (n=36/38, %)** |
| --- | --- | --- | --- |
| **Incorrect folder names** | B_COAGDISAESI_Coagulation disorders all, BAIHEMOLYTICANEMIA_AESI_Autoimmune hemolytic anemia | Module 1, check #1 (S4) | 2/2 (100) |
| **Missing or multiple files in several folders** | C_ACI_AESI_Acute cardiovascular injury, C_AMI_AESI_Acute myocardial Infarction, C_ANGINA_AESI_Angina pectoris | Module 1, Check #2 | 3/3 (100) |
| **Excel filename does not match folder concept name** | Excel name not matching variable name (from folder name) B_ANAEMIA_COV_Anaemia | Module 1, check #5 | 1/1 (100) |
| **Worksheet name does not match Excel file name** | Worksheet name not matching Excel file name C_ACUTEHF_AESI_Acute heart failure | Module 1, check #6 | 1/1 (100) |
| **Concepts and algorithms mismatch** | Algorithm references non-existent concepts folders available (Table 3 in Supplemental File 2) | Module 1 Check #7 | 21/21 (100) |
| **Metadata mismatches in library metadata** | Event listed in library metadata but not present in concept folder (Table 4 in Supplemental file 2) | Module 2, Check 4 | 1/3 (33) |
| **Missing required concept codelist for the study** | Concept folders and Algorithms selected for the study but not present in the output codelist (Table 3 and Table 5 in Supplemental file 2) | Module 2, Check #2 | 26/26 (100) |
| **Missing values (e.g., missing codes, tags)** | Missing event code | Module 2, Check #7 | 4/4 (100) |
| **Code formatting issue due to rounding** | Codes longer than 15 digits ending in 3 zeros | Module 2, Check #8 | 2/2 (100) |

## 4. DISCUSSION

The increasing use of RWD demands efficient and reproducible approaches for extracting clinical concepts through codelists. The development of CodeMergeR directly addresses the challenge of concatenating individual codelists through two integrated modules, offering a quality assurance solution for codelist concatenation.

### 4.1 Key Findings

CodeMergeR can reliably identify a wide range of formatting, structural, and semantic errors. For example, the code 1240531000000103 (Myocarditis due to disease caused by severe acute respiratory syndrome coronavirus 2 (disorder) in SNOMED vocabulary is rounded when used in Excel (i.e., the last 103 digits become 000). The findings in Module Conformance stress the importance of robust file structures and careful data formatting in the development and management of codelists. For example, the detection of multiple files within a single folder and the mismatch in file names illustrate how seemingly minor inconsistencies can disrupt automated workflows. Similarly, issues identified in Module cleaning and standardization such as missing entries in the concept directory, structural anomalies, and warnings related to potential rounding of codes, highlight the value of automated validation tools. These types of errors would be difficult to detect manually in a timely and reliable manner, reinforcing the need for systematic quality checks during codelist preparation.

Organizing CDM such as ConcePTION (i.e., no semantic harmonization is required) generally require a broader set of quality checks than rule-based CDMs like OMOP.^11,27^ Because the ConcePTION CDM allows multiple vocabularies, a summary of the all included vocabularies is necessary to detect missing or unregistered ones, handle range-style values and reconcile conflicting tags in clinical concepts (i.e., the same code appears as high-specificity and low-specificity) . In contrast, OMOP requires data holders to map the original codes to a common vocabulary (SNOMED CT) before analysis. While errors in this mapping process are not always transparent, OMOP does not apply cross-vocabulary reconciliation during analytical stage as ConcePTION CDM does.^28^

The internal validation process, involving 38 manually introduced errors, confirmed an overall detection rate of 96.8%, with all major error categories such as naming convention issues, missing files, and erroneous codes successfully identified. This process is critical in the context of large-scale RWE studies where even minor inconsistencies can lead to the exclusion of key clinical concepts or misclassification of outcomes.

### 4.2. Strength and Limitations

CodeMergeR processed individual concept codelist files including 50 clinical concepts folders and 10 algorithms within one minute, with consistent performance across operating systems. Its user-friendly interface, including instructional alerts, clickable legends, and structured outputs, supports adoption by non-technical users such as clinicians and researchers. The use of CodeMergeR within VAC4EU and EU PE&PV highlights its value for collaborative research environments, where codelists are iteratively reviewed, shared, and reused across studies. By centralizing and standardizing codelist processing, the application reduces duplication of effort, saves time, provides documentation on issues or errors, and enhances reproducibility.

The current version of CodeMergeR is tightly coupled to a particular folder structure. Its open-source license allows for adaptation to other international networks that need to deal with concept codelist concatenation. Integration with external code repositories or public-facing platforms like OpenCodelists could further expand its utility and ensure alignment with evolving standards in the field.^29^ Additionally, the validation process does not yet include comprehensive checks of the library metadata content, as reflected in a 33% detection rate for metadata-related issues. This app was initially created to check and harmonize individual codelists into a unified master file, but further enhancements should be added for issues such as misspelling of clinical concepts which might lead to missingness of necessary clinical concepts in the master codelist.

### 4.3 Comparison with other tools

Several tools exist to support the creation and management of codelists, each supporting different stages of the RWE workflow. DARWIN EU’s CodelistGenerator is a data-driven R package designed to work within the OMOP CDM.^27,30^ It connects directly to OMOP data instances and returns candidate codes using keywords/regex and options such as ‘includeDescendants’, offering diagnostics to assess codes appearance in the data. Its primary function is to assist researchers in discovering and defining clinical concepts based on observed data. CodeMergeR complements CodelistGenerator by operating downstream in the workflow. While CodelistGenerator helps identify relevant codes, CodeMergeR focuses on standardizing and validating researcher-refined codelists, often compiled from multiple sources and vocabularies into a harmonized master file. Together, these tools could form a pipeline: discovery with CodelistGenerator, followed by quality assurance and integration with CodeMergeR.

OHDSI ATHENA serves as a vocabulary browser and downloader for OMOP-standardized vocabularies, but lacks functionality for integrating or cleaning externally sourced, or multi-vocabulary codelists.^31^ CodeMergeR could bridge this gap by transforming heterogeneous codelists into OMOP-compatible formats, enabling smoother downstream use in tools like ATLAS.^32^ OpenCodelists, developed by OpenSAFELY, provides a collaborative web interface for building, reviewing, versioning, and sharing codelists.^29^ While it supports transparency and reuse, it lacks automated harmonization features. CodeMergeR could serve as a pre-processing step before uploading to OpenCodelists, ensuring that lists are clean and structurally sound for the platform.

### 4.4 Disclaimer

Given the lessons learned during the development of *CodeMergeR,* VAC4EU has improved CodeMapper leading to a version 2.0 (manuscript in preparation) which integrates all steps of codelist creation, tagging, and concatenation.

## 5. CONCLUSIONS

CodeMergeR is an open-source R Shiny application developed to streamline the processing of codelists for RWE studies. It automates the standardization, cleaning, and integration of files containing clinical codes from multiple vocabularies that have been collaborative and iteratively reviewed. By enforcing metadata standards and automating validation checks, the tool improves reproducibility, minimizes manual effort, reduces workload, and supports scalable, transparent codelist management. Future development will focus on expanding compatibility with alternative metadata formats, folder system structures, incorporating user feedback and integrating CodeMergeR with public tools and repositories to enhance interoperability and broader adoption, improving the codelist lifecycle.

## FUNDING STATEMENT

No external funding was received for the development of this project.

## AUTHORS CONTRIBUTION

VH and CLAN started with the initial design; V.H programmed the R scripts; VH and CLAN wrote the initial draft of the manuscript. VH, CLAN, JRA, SM, and MCJMS provided critical feedback and edited the manuscript. This paper only reflects the personal views of the stated authors.

## Data Availability

All data produced in the present work are contained in the manuscript

## ACKNOWLEDGMENTS

We would like to thank VAC4EU codelist taskforce (Carlos E Durán, Cristina Rebordosa, Joan Fortuny, Alejandro Arana, Vera Ehrenstein, Andrea Margulis, Samantha Lane, Denise Morris, Taylor Aurelius, Benedikt Becker) for supporting codelist creation, review, and maintenance of the VAC4EU library and the VAC4EU Principal Investigators for elevating requirements and testing the tool.

## CONFLICT OF INTEREST STATEMENT

VH, CLAN, JRA, SM, and MCJMS are currently salaried employees at University Medical Center Utrecht, which receives institutional research funding from pharmaceutical companies and regulatory agencies and is administered by University Medical Center Utrecht.

## CodeMergeR AVAILABILITY

CodeMergeR is available as an open-source R scripts in the following link https://github.com/vjolahoxhaj/CodeMergeR/. The repository has a MIT License (NonCommercial).

## ETHICS STATEMENT

The authors state that no ethical approval was needed.

## SUPPLEMENTARY MATERIAL

**Supplementary file 1.** Examples of the input files for CodeMergeR

**Supplementary file 2.** Description of the errors introduced to the codelist to test app performance

**Supplementary file 3.** File containing the supplemental figures

## DECLARATION OF GENERATIVE AI IN SCIENTIFIC WRITING

During the preparation of this work, the author(s) used GTP-4 to improve readability and language. After using this tool, the author reviewed and edited the content as needed and take full responsibility for the content.

## Notes

### Competing Interest Statement

The authors have declared no competing interest.

## REFERENCES

1. Dang A. Real-World Evidence: A Primer. Pharmaceut Med. Jan 2023;37(1):25–36. doi:10.1007/s40290-022-00456-6

2. Casey JA, Schwartz BS, Stewart WF, Adler NE. Using Electronic Health Records for Population Health Research: A Review of Methods and Applications. Annu Rev Public Health. 2016;37:61–81. doi:10.1146/annurev-publhealth-032315-021353

3. Liu F, Panagiotakos D. Real-world data: a brief review of the methods, applications, challenges and opportunities. BMC Med Res Methodol. Nov 5 2022;22(1):287. doi:10.1186/s12874-022-01768-6

4. NHS Digital. Read codes. Accessed 18 October, 2025. https://digital.nhs.uk/services/terminology-and-classifications/read-codes

6. WONCA International Classification Committee. International classification of primary care, version 2 (ICPC-2). Accessed 18 October, 2025. https://www.who.int/standards/classifications/other-classifications/international-classification-of-primary-care

6. SNOMED International. SNOMED CT: The global language of healthcare. Accessed 18 October, 2025. https://www.snomed.org

7. World Health Organization (WHO). International classification of diseases (ICD). Accessed 18 October, 2025. https://www.who.int/classifications/icd/en/

8. Nederlands Huisartsen Genootschap. NHG-Tabel-78 ICPC-1 NL-Engels-versie 1-Inkijkexemplaar. Accessed 18 October, 2025. https://referentiemodel.nhg.org/sites/default/files/public/NHG-Tabel%2078-ICPC-1%20NL-Engels-versie%201-Inkijkexemplaar.pdf

9. BioPortal. SNOMED CT: Guillain-Barré syndrome. Accessed 18 October, 2025. https://bioportal.bioontology.org/ontologies/SNOMEDCT?p=classes&conceptid=40956001

10. Matthewman J, Andresen K, Suffel A, et al. Checklist and guidance on creating codelists for routinely collected health data research. NIHR Open Res. 2024;4:20. doi:10.3310/nihropenres.13550.2

11. Thurin NH, Pajouheshnia R, Roberto G, et al. From Inception to ConcePTION: Genesis of a Network to Support Better Monitoring and Communication of Medication Safety During Pregnancy and Breastfeeding. Clin Pharmacol Ther. Jan 2022;111(1):321–331. doi:10.1002/cpt.2476

12. Becker BFH, Avillach P, Romio S, et al. CodeMapper: semiautomatic coding of case definitions. A contribution from the ADVANCE project. Pharmacoepidemiol Drug Saf. Aug 2017;26(8):998–1005. doi:10.1002/pds.4245

13. Gini R. Report with publications on results of component algorithms (D7.16). 2024.

14. R: A language and environment for statistical computing. R Foundation for Statistical Computing; 2021. https://www.R-project.org/

15. stringr: Simple, Consistent Wrappers for Common String Operations. *R package version 1.6.0. 2025.* https://CRAN.R-project.org/package=stringr

16. data.table: Extension of ‘data.fram’. R package version 1.17.8. 2025. https://CRAN.R-project.org/package=data.table

16. knitr: A General-Purpose Package for Dynamic Report Generation in R. R package version 1.50. 2025. https://yihui.org/knitr/

18. Xie Y CJ, Tan X. DT: A Wrapper of the JavaScript Library ’DataTables’. R package version 0.33. 2024;

19. shiny: Web Application Framework for R. R package version 1.10.0. 2024. https://CRAN.R-project.org/package=shiny

20. shinyjs: Easily Improve the User Experience of Your Shiny Apps in Seconds. *R package version 2.1.0*. 2021. https://CRAN.R-project.org/package=shinyjs

21. rstudioapi: Safely Access the RStudioAPI. R package version 0.17.1. 2024. https://CRAN.R-project.org/package=rstudioapi

22. readxl: Read Excel Files. R package version 1.4.5. 2025. https://CRAN.R-project.org/package=readxl

23. htmltools: Tools for HTML. R package version 0.5.8.1. 2024. https://CRAN.R-project.org/package=htmltools

24. tinytex: Helper Functions to Install and Maintain TeX Live, and Compile LaTeX Documents. R package version 0.57. 2025. https://github.com/rstudio/tinytex

25. shinyscreenshot: Capture Screenshots of Entire Pages or Parts of Pages in ’Shiny’. R package version 0.2.1. 2023. https://CRAN.R-project.org/package=shinyscreenshot

26. rmarkdown: Dynamic Documents for R. R package version 2.30. 2025. https://github.com/rstudio/rmarkdown

27. Observational Health Data Sciences and Informatics (OHDSI). Data Standardization – OHDSI. Accessed 18 October, 2025. https://www.ohdsi.org/data-standardization/

28. Reich C, Ostropolets A, Ryan P, et al. OHDSI Standardized Vocabularies-a large-scale centralized reference ontology for international data harmonization. J Am Med Inform Assoc. Feb 16 2024;31(3):583–590. doi:10.1093/jamia/ocad247

29. OpenSAFELY. OpenCodelists. Accessed 18 October, 2025. https://www.opencodelists.org/

30. DARWIN EU. CodelistGenerator: Identify Relevant Clinical Codes and Evaluate Their Use. Accessed 18 October, 2025. https://darwin-eu.github.io/CodelistGenerator/?utm_source=chatgpt.com

31. Observational Health Data Sciences and Informatics (OHDSI). Athena: Search Terms. Accessed 18 October, 2025. https://athena.ohdsi.org/search-terms/terms

32. Observational Health Data Sciences and Informatics (OHDSI). ATLAS: OHDSI Demo. Accessed 18 October, 2025. https://atlas-demo.ohdsi.org/

