## Supplemental file 3 for "CodeMergeR: A shiny-based application for codelist integration"

**Functional Testing**

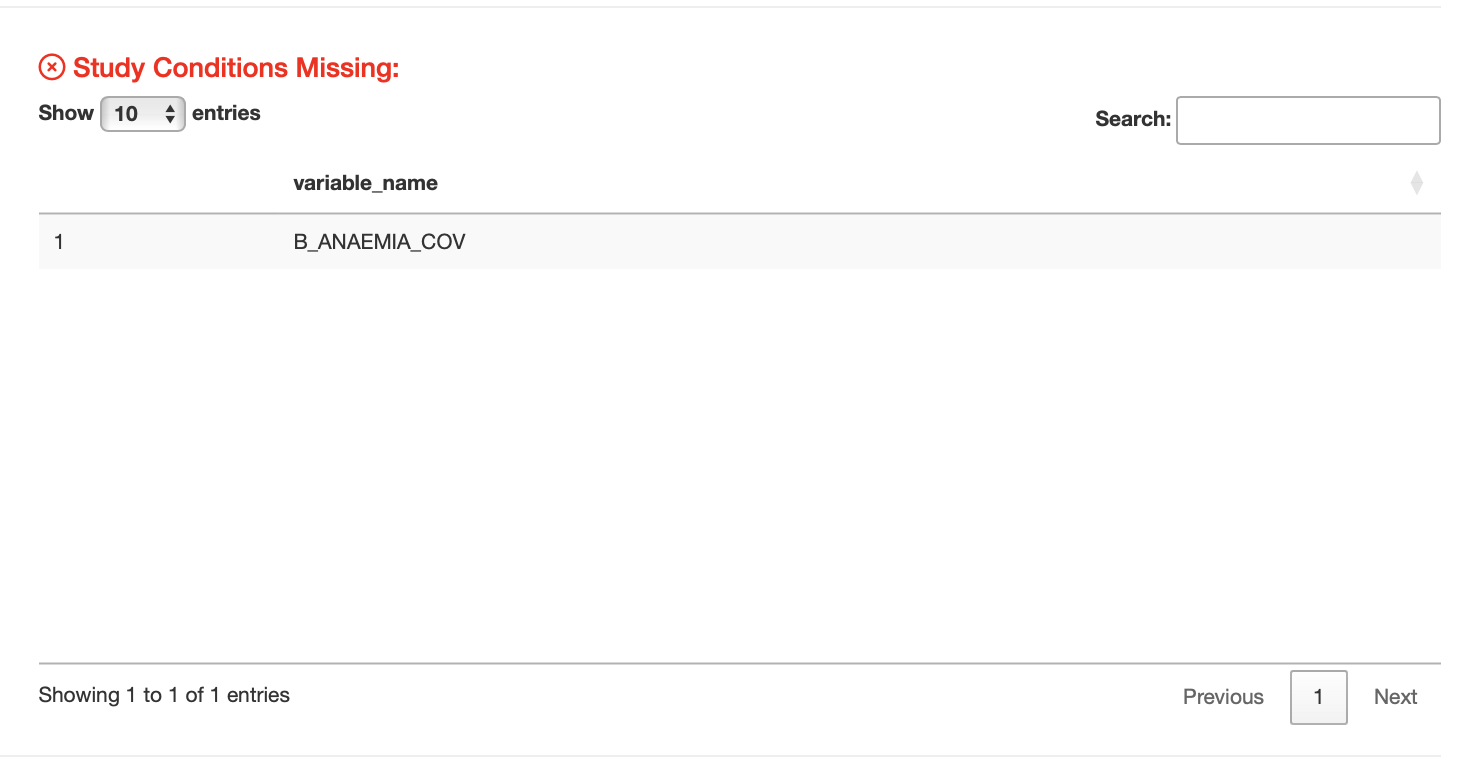

Figure S1. Visualization module 2 check #1

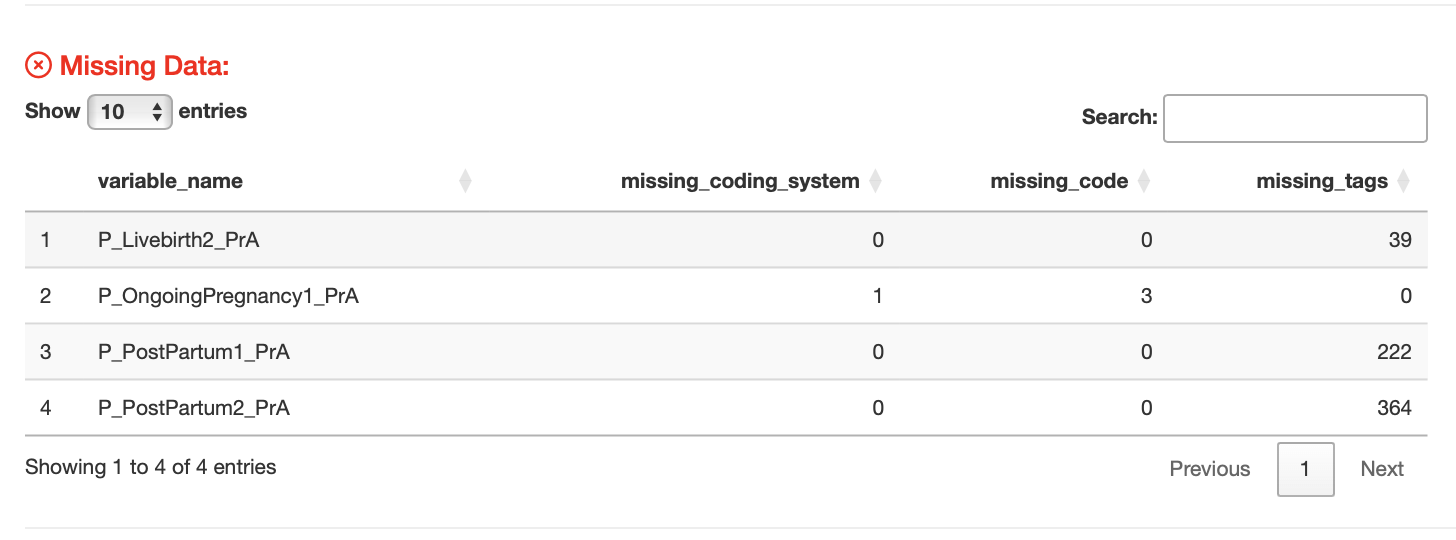

Figure S2. Visualization module 2 check #7

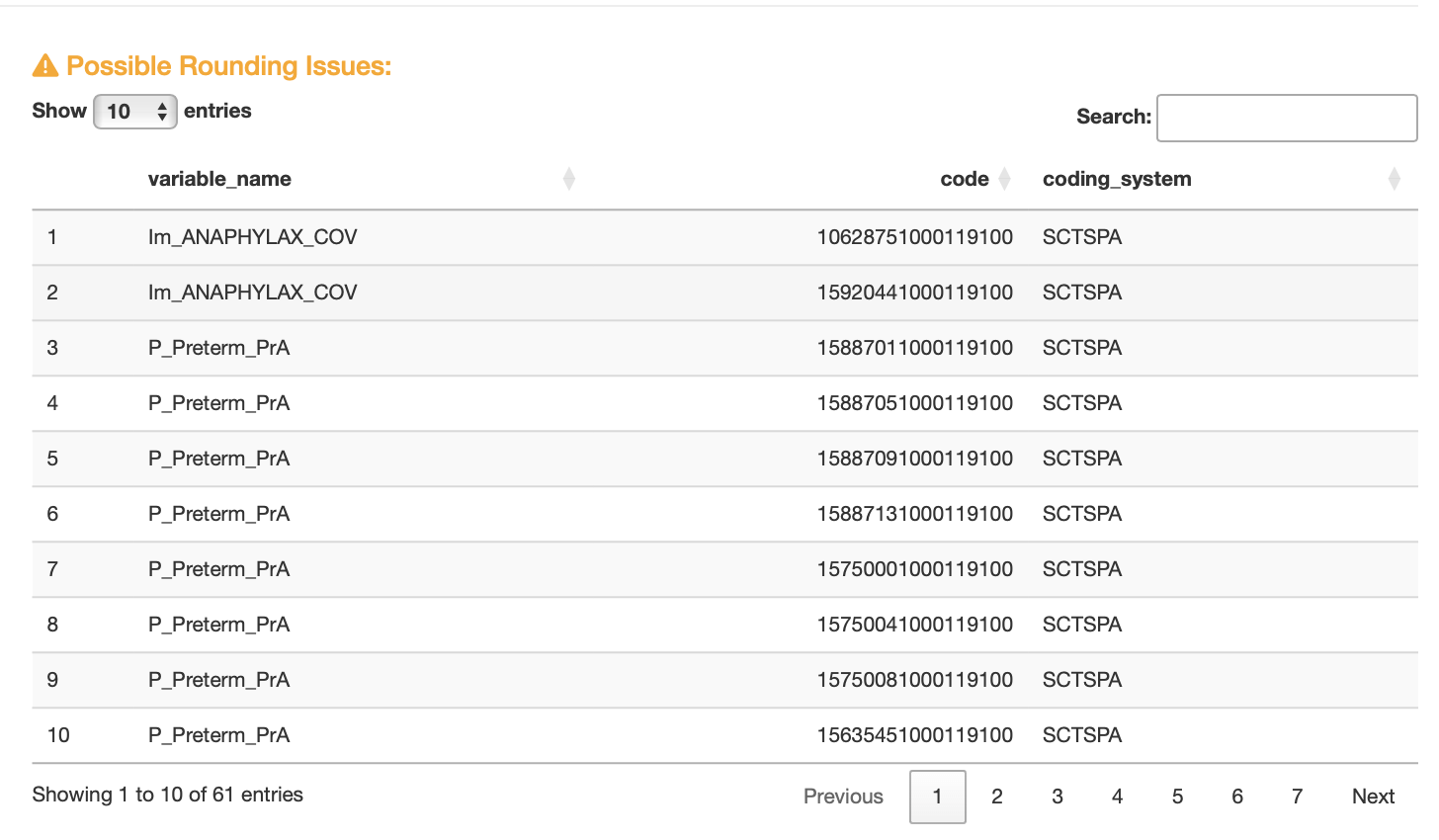

Figure S3. Visualization module 2 check #9

**Performance testing**

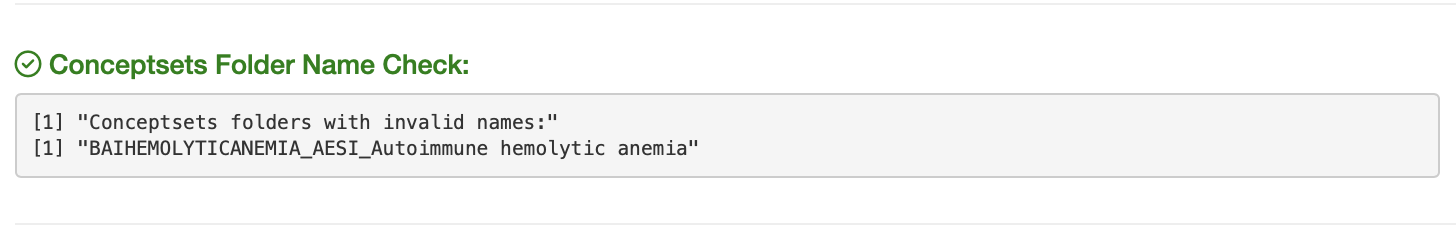

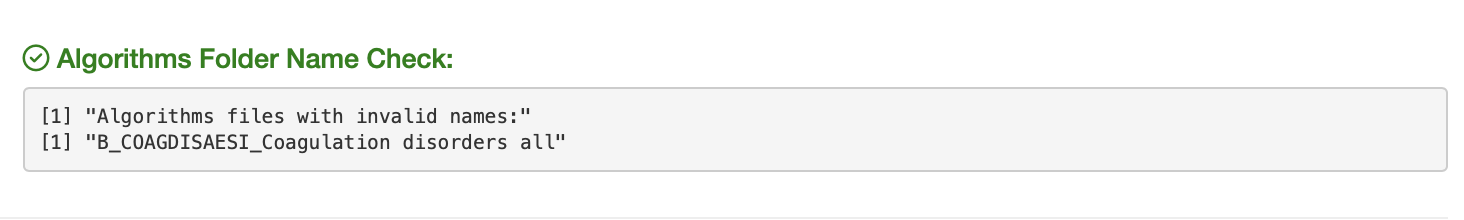

Figure S4. Visualization of Incorrect folder names
