## Supplemental file 2 for "CodeMergeR: A shiny-based application for codelist integration"

TableS 1: List of Algorithms’ folders and manually introduced errors

| **Algorithms** |  |  |
| --- | --- | --- |
| **Algorithm** | **Error Algorithm** | **Introduced error** |
| B_COAGDIS_AESI_Coagulation  disorders all | B_COAGDISAESI_Coagulation disorders all | Naming convention error – removal of one underscore  Extra excel file inside folder |
| C_ACI_AESI_Acute cardiovascular injury |  | Double .xls files |
| C_CARDIOCEREBROVASCULARDESE_COV_CardioCerebroVascular disease |  |  |
| D_LIVERCHRONIC_COV_Liver chronic disease |  |  |
| E_DM12ALGORITHM_COV_Diabetes 1 and 2 |  |  |
| I_HIV_COV_HIV |  |  |
| I_RESPINFECALGORITHM_COV_Respiratory infection algorithm |  |  |
| Im_ALLERGY_COV_Allergy |  |  |
| Im_ANAPHYLAXIS_COV_Anaphylaxis history |  |  |
| L_OBESITYALGORITHM_COV_Obesity |  |  |

Table S2: List of available COIs folders and manually introduced errors

| **Concept folders** |  |  |
| --- | --- | --- |
| **Concepts** | **Error concepts** | **Introduced error** |
| B_AIHEMOLYTICANEMIA_AESI_Autoimmune hemolytic anemia | BAIHEMOLYTICANEMIA_AESI_Autoimmune hemolytic anemia | Naming convention error – removal of one underscore |
| B_ANAEMIA_COV_Anaemia |  | The name of the excel file doesn’t match with the variable name retrieved from the folder name (B_ANEM_COV.xlsx) |
| C_ACUTEHF_AESI_Acute heart failure |  | The name of the worksheet doesn’t match with the name of the excel file (C_ACUTE_AESI) |
| C_AMI_AESI_Acute myocardial Infarction |  | No excel file |
| C_ANGINA_AESI_Angina pectoris |  | No excel file |
| C_SCD_AESI_Death sudden cardiac |  |  |
| C_SHOCKCARD_AESI_Shock Cardiogenic |  |  |
| C_VALVULAR_AESI_Valvular heart disease non-congenital not rheumatic |  |  |
| D_GALLSTONES_COV_Gall stones |  |  |
| D_GERD_AESI_Gastroesophageal reflux disease |  |  |
| D_HEPIMPAIRMENT_COV_Hepatic impairment unspecified for being mild or moderate or sever |  |  |
| D_IBD_COV_Inflammatory bowel disease ulcerative colitis and Crohns disease |  |  |
| E_DIABETESNOCOMPLICATIONS_COV_Diabetes without chronic complication |  |  |
| E_DM1_AESI_Diabetes type 1 |  |  |
| E_DM2_COV_Diabetes mellitus type 2 |  |  |
| E_DMCOMPL_COV_Chronic Diabetes complications |  |  |
| E_GOITER_AESI_Goiter |  |  |
| E_GOUT_AESI_Gout |  |  |
| I_AIDS_COV_AIDS Charlson index |  |  |
| I_BACTERIALINFEC_AESI_Bacterial infections |  |  |
| I_CAMPYLOBACTERJEJ_COV_Campylobacter jejuni infection |  |  |
| I_CHICKENPOX_COV_Chickenpox |  |  |
| I_CHLAMYDIA_COV_Chlamydia |  |  |
| I_CMV_COV_Cytomegalovirus infection excluding congenital ones |  |  |
| I_COVID19DX_AESI_COVID19 diagnosis outcome |  |  |
| I_COVID19DX_COV_COVID19 diagnosis |  |  |
| I_KLEBSIELLA_COV_Klebsiella infection |  |  |
| I_MALARIA_COV_Malaria |  |  |
| L_OBESE_COV_Obese |  |  |
| L_OBESITY_COV_Obesity diagnosis |  |  |
| L_OVERWEIGHT_COV_Overweight |  |  |
| M_FRACTURES_AESI_Fractures non-osteoporotic fractures |  |  |
| M_FRAGILITYFRACTURE_COV_Fragility fracture |  |  |
| M_HIPFRACTURE_COV_Hip fracture |  |  |
| M_RPM_COV_Polymyalgia rheumatica |  |  |
| Ment_ADHD_COV_Attention deficit hyperactivity disorder |  |  |
| Ment_ALCABUSE_COV_Alcohol abuse |  |  |
| Ment_ANXIETY_COV_anxiety |  |  |
| N_VERTIG_COV_Vertigo |  |  |
| O_FLARE_AESI_Unspecific disease flare up |  |  |
| O_LEARNDISAB_COV_Learning disability |  |  |
| P_24weeksCHILD_PrA_24 weeks pregnancy CHILD |  |  |
| P_24weeksLB_PrA_24 weeks pregnancy |  |  |
| P_24weeksUNK_PrA_24 weeks pregnancy unknown |  |  |
| P_APGARLOW_AESI_Low 5min Apgar |  |  |
| P_AtTerm_PrA_At term delivery |  |  |
| P_AtTermLB_PrA_At term delivery with live birth |  |  |
| P_BirthChild_PrA_Birth of a child |  |  |
| P_OngoingPregnancy6_PrA_Ongoing Pregnancy |  |  |
| V_VASCULITISANY_COV_Vasculitis any |  |  |

Table S3: List of individual concepts necessary for the available Algorithms

| **Necessary COIs for Algorithms construction** |  |  |
| --- | --- | --- |
| **Algorithm** | **Variable_name** | **Presence in test folder** |
| C_CARDIOCEREBROVASCULARDESE_COV | C_AMI_AESI | Yes, but previous error |
| C_CARDIOCEREBROVASCULARDESE_COV | C_ANGINA_AESI | Yes, but previous error |
| C_CARDIOCEREBROVASCULARDESE_COV | C_CARDIOMYOPATHY_COV | No |
| C_CARDIOCEREBROVASCULARDESE_COV | C_HF_AESI | No |
| C_CARDIOCEREBROVASCULARDESE_COV | C_MYOCARD_AESI | No |
| C_CARDIOCEREBROVASCULARDESE_COV | C_MYOPERICARD_AESI | No |
| C_CARDIOCEREBROVASCULARDESE_COV | C_PERICARD_AESI | No |
| D_LIVERCHRONIC_COV | D_ALCOHOLICLIVER_COV | No |
| D_LIVERCHRONIC_COV | D_HEPATITISAUTOIMMUNE_COV | No |
| D_LIVERCHRONIC_COV | D_LIVERCHRONICALONE_COV | No |
| D_LIVERCHRONIC_COV | D_LIVERCIRRHOSIS_AESI | No |
| D_LIVERCHRONIC_COV | D_NONALCOHOLICLIVER_COV | No |
| E_DM12ALGORITHM_COV | E_DM12_COV | No |
| I_HIV_COV | I_AIDS_COV | Yes |
| I_RESPINFECALGORITHM_COV | I_INFLUENZA_AESI | No |
| I_RESPINFECALGORITHM_COV | I_RESPINFEC_COV | No |
| Im_ALLERGY_COV/ Im_ANAPHYLAXIS_COV | Im_ANAPHYLAXIS_AESI | No |
| Im_ALLERGY_COV | Im_HYPERSENS_AESI | No |
| L_OBESITYALGORITHM_COV | L_OBESITY_COV | Yes |
| L_OBESITYALGORITHM_COV | L_OVERWEIGHT_COV | Yes |
| C_CARDIOCEREBROVASCULARDESE_COV | N_STROKEHEMO_AESI | No |
| C_CARDIOCEREBROVASCULARDESE_COV | N_STROKEISCH_AESI | No |
| C_CARDIOCEREBROVASCULARDESE_COV | N_TIA_AESI | No |
| C_CARDIOCEREBROVASCULARDESE_COV | V_ANEURYSMVASCMALF_COV | No |

Table S4: library metadata file and manually introduced errors

| **Event_abbreviation / Variable Name** | **[Study_Name]** | **Introduced error** |
| --- | --- | --- |
| E_GOITER_AESI | N/A | Missing in  **Event_abbreviation / Variable Name** |
| B_DIC_AESI | No | Extra in  **Event_abbreviation / Variable Name,** missing in concept folders |
| D_VOMITING_AESI | Yes | Extra in  **Event_abbreviation / Variable Name,** missing in concept folders |

Table S5: library metadata file (the first 4 columns are to be found in the Codelist library metadata Excel file, the last column display whether this an error or not)

| **Event_abbreviation / Variable Name** | **Events & subconcepts for diagnosis codes** | **Type of event** | **[Study_Name]** | **Presence in test folders** |
| --- | --- | --- | --- | --- |
| B_ANAEMIA_COV | Anaemia | COV | yes | Yes, but previous error |
| B_AIHEMOLYTICANEMIA_AESI | Autoimmune hemolytic anemia | AESI | yes | Yes, but previous error |
| B_COAGDEF_AESI | Coagulation, deficiencies | AESI | yes | No |
| B_DIC_AESI | Coagulation, disseminated intravascular | AESI |  | No |
| C_ACI_AESI | Acute cardiovascular injury | AESI | yes | Yes, but previous error |
| C_ACUTEHF_AESI | Acute heart failure | AESI | yes | Yes, but previous error |
| C_AMI_AESI | Infarction, myocardial | AESI | yes | Yes, but previous error |
| C_ANGINA_AESI | Angina pectoris | AESI | yes | Yes, but previous error |
| C_CARDIOCEREBROVASCULARDESE_COV | CardioCerebroVascular disease | COV |  | Yes, Algorithm |
| C_SCD_AESI | Death, sudden cardiac | AESI | yes | Yes |
| C_SHOCKCARD_AESI | Shock, cardiogenic | AESI | yes | Yes |
| C_VALVULAR_AESI | Valvular heart disease (non-congenital, not rheumatic) | AESI |  | Yes |
| D_GALLSTONES_COV | Gall stones | COV |  | Yes |
| D_IBD_COV | Inflammatory bowel disease (ulcerative colitis and Crohn’s disease) | COV | yes | Yes |
| D_LIVERCHRONIC_COV | Liver chronic disease | COV | yes | Yes, Algorithm |
| D_VOMITING_AESI | Vomiting as general symptoms and not those associated with specific diseases such as those related to pregnancy, or in the newborn, following surgery or after surgery. | AESI | yes | No |
| D_HEPIMPAIRMENT_COV | Hepatic impairment unspecified for being mild or moderate or sever | COV |  | Yes |
| D_GERD_AESI | Gastroesophageal reflux disease | AESI |  | Yes |
| E_DIABETESNOCOMPLICATIONS_COV | Diabetes without chronic complication | COV | yes | Yes |
| E_DM1_AESI | Diabetes (type 1) | AESI | yes | Yes |
| E_DM12ALGORITHM_COV | Diabetes (types 1 and 2) | COV | yes | Yes, Algorithm |
| E_DM2_COV | Diabetes (type 2) | COV |  | Yes |
| E_GOUT_AESI | Gout | AESI |  | Yes |
| E_GOITER_AESI | Goiter | AESI |  | Yes |
| I_AIDS_COV | AIDS | COV | yes | Yes |
| I_COVID19DX_AESI | COVID-19, diagnosis outcome | AESI | yes | Yes |
| I_COVID19DX_COV | COVID‑19, diagnosis | COV | Yes | Yes |
| I_HIV_COV | HIV | COV | yes | Yes, Algorithm |
| **I_RESPINFECALGORITHM_COV** | **Respiratory infection algorithm** | COV | Yes | Yes |
| I_CMV_COV | **Cytomegalovirus excluding congenital ones** | COV | yes | Yes |
| I_BACTERIALINFEC_AESI | Bacterial infections | AESI | yes | Yes |
| I_CHLAMYDIA_COV | Chlamydia | COV | yes | Yes |
| I_MALARIA_COV | Malaria | COV | yes | Yes |
| Im_ALLERGY_COV | Allergy | COV | yes | Yes, Algorithm |
| Im_ANAPHYLAX_COV | Anaphylaxis history (Component of anaphylaxis history algorithm ) | COV | yes | No, Algorithm |
| L_OBESITY_COV | Obesity | COV | yes | Yes |
| L_OBESE_COV | Obese | COV | yes | Yes |
| **L_OBESITYALGORITHM_COV** | **Obesity** | COV | yes | Yes, Algorithm |
| L_OVERWEIGHT_COV | Overweight | COV | yes | Yes |
| M_FRACTURES_AESI | Fractures non-osteoporotic and osteoporotic | AESI | yes | Yes |
| M_FRAGILITYFRACTURE_COV | Fragility fracture | COV | yes | Yes |
| M_HIPFRACTURE_COV | Hip fracture | COV |  | Yes |
| M_RPM_COV | Polymyalgia rheumatica | COV | yes | Yes |
| Ment_ADHD_COV | Attention-deficit/hyperactivity disorder (ADHD) | COV | yes | Yes |
| Ment_ALCABUSE_COV | Alcohol, abuse | COV | yes | Yes |
| Ment_ANXIETY_COV | Anxiety | COV |  | Yes |
| N_VERTIG_COV | Vertigo | COV | yes | Yes |
| O_LEARNDISAB_COV | Learning disability | COV | yes | Yes |
| O_FLARE_AESI | Unspecific disease flare up | AESI | yes | Yes |
| P_24weeksLB_PrA | Gestation 24 weeks LB | PrA | yes | Yes |
| P_24weeksCHILD_PrA | Gestation 24 weeks CHILD | PrA | yes | Yes |
| P_24weeksUNK_PrA | Gestation 24 weeks unknown | PrA | yes | Yes |
| P_AtTerm_PrA | At term delivery | PrA | yes | Yes |
| P_APGARLOW_AESI | Pregnancy, Low 5-minute Apgar score | AESI | yes | Yes |
| P_BirthChild_PrA | Birth of a Child | PrA | yes | Yes |
| P_OngoingPregnancy6_PrA | Ongoing pregnancy | PrA | yes | Yes |
| P_AtTermLB_PrA | At term delivery with live birth | PrA | yes | Yes |
| V_VASCULITISANY_COV | Vasculitis, any | COV | yes | Yes |
| I_KLEBSIELLA_COV | Klebsiella infection | COV | yes | Yes |
| I_CAMPYLOBACTERJEJ_COV | Campylobacter jejuni infection | COV | yes | Yes |
| I_CHICKENPOX_COV | Chickenpox | COV | yes | Yes |
