## Supplemental file 1 for "CodeMergeR: A shiny-based application for codelist integration"

Table S1: Algorithms’ Excel file structure

**Algorithm:** B_COAGDIS_AESI_Coagulation disorders all

**Codesheet_name**

B_DIC_AESI

B_ITP_AESI

B_TP_AESI

B_VITT_AESI

B_SPLACHNICVT_AESI

N_CVST_AESI

R_PE_AESI

V_DVT_AESI

V_THROMBOSISARTERIAL_AESI

V_OTHERVTE_AESI

B_COAGDEF_AESI

Table S2: COIs’ Excel file structure

| **COI:** B_COAGDEF_AESI_Coagulation deficiencies | | | | | |
| --- | --- | --- | --- | --- | --- |
| **Coding_system** | **Code** | **Code name** | **Concept** | **Concept name** | **tags** |
| ICD10CM | D65 | Diffuse or disseminated intravascular coagulation [DIC] | C2873782 | Diffuse or disseminated intravascular coagulation [DIC] | narrow |
| ICD10CM | D65 | Disseminated intravascular coagulation [defibrination syndrome] | C0012739 | Disseminated Intravascular Coagulation | narrow |
| ICD10CM | D66 | Hereditary factor VIII deficiency | C0019069 | Hemophilia A | exclude |
| SNOMEDCT_US | 11093006 | Angiohemophilia | C0042974 | von Willebrand Disease | possible |
| SNOMEDCT_US | 191287000 | Hemorrhagic disorder due to circulating anticoagulants | C1399404 | Hemorrhagic disorder due to circulating anticoagulants | narrow |
| SNOMEDCT_US | 191289002 | Hemorrhagic disorder due to hyperheparinemia | C0272273 | Hemorrhagic disorder due to hyperheparinemia | narrow |

Table S3: Codelist library metadata file structure

| **General Clinical Concept** | **Events Relationship Grouping Tag** | **Event_abbreviation / Variable Name** | **Events & subconcepts for diagnosis codes** | **Type of event** | **Covariates with no codes** | **Test_data** |
| --- | --- | --- | --- | --- | --- | --- |
| B - Blood and hematopoietic organs diseases | Part of B_COAGDIS_AESI | B_COAGDEF_AESI | Coagulation, deficiencies | AESI |  |  |
| B - Blood and hematopoietic organs diseases | Part of frailty | B_COAGDEF_COV | Coagulation, deficiencies | COV |  | **yes** |
| B - Blood and hematopoietic organs diseases |  | B_COAGDIS_AESI | Coagulation disorders, all | AESI | yes |  |
| D - Digestive tract diseases | PART OF: D_LIVERCHRONIC_COV | D_LIVERCHRONICALONE_COV | Liver chronic disease only | COV |  |  |
| D - Digestive tract diseases | part of D_DIGESTIVEDIS_AESI algorithm | D_ULCERATIVECOLITIS_AESI | Ulcerative colitis | AESI |  |  |
| I - Infectious diseases | PART OF: I_HIV_COV | I_AIDS_COV | AIDS | COV |  |  |
| I - Infectious diseases | Part of P_MATERNALTORCH_COV algorithm | I_ZIKA_COV | Zika virus | COV | yes |  |
